# Modelling testing and response strategies for COVID-19 outbreaks in remote Australian Aboriginal communities

**DOI:** 10.1101/2020.10.07.20208819

**Authors:** Ben B. Hui, Damien Brown, Rebecca H. Chisholm, Nicholas Geard, Jodie McVernon, David G. Regan

**Affiliations:** Kirby Institute, UNSW Sydney, Sydney, NSW, Australia; The Peter Doherty Institute for Infection and Immunity, The Royal Melbourne Hospital and The University of Melbourne, Australia; Centre for Epidemiology and Biostatistics, Melbourne School of Population and Global Health, The University of Melbourne, Australia; Department of Mathematics and Statistics, School of Engineering and Mathematical Sciences, La Trobe University, Australia; Centre of Epidemiology and Biostatistics, Melbourne School of Population and Global Health, University of Melbourne, Australia; School of Computing and Information Systems, The University of Melbourne, Melbourne, Australia; Victorian Infectious Diseases Reference Laboratory Epidemiology Unit, The Peter Doherty Institute for Infection and Immunity, The Royal Melbourne Hospital and The University of Melbourne, Australia

**Keywords:** COVID-19, Indigenous health, Outbreaks, Quarantine, Patient Isolation, Households, Family and Household

## Abstract

**Background:** Remote Australian Aboriginal and Torres Strait Islander communities have potential to be severely impacted by COVID-19, with multiple factors predisposing to increased transmission and disease severity. Our modelling aims to inform optimal public health responses.

**Methods:** An individual-based simulation model represented communities ranging from 100 to 3,500 people, comprised of large interconnected households. A range of strategies for case finding, quarantining of contacts, testing, and lockdown were examined, following the silent introduction of a case.

**Results:** Multiple secondary infections are likely present by the time the first case is identified. Quarantine of close contacts, defined by extended household membership, can reduce peak infection prevalence from 60-70% to around 10%, but subsequent waves may occur when community mixing resumes. Exit testing significantly reduces ongoing transmission.

Concurrent lockdown of non-quarantined households for 14 days is highly effective for epidemic control and reduces overall testing requirements; peak prevalence of the initial outbreak can be constrained to less than 5%, and the final community attack rate to less than 10% in modelled scenarios. Lockdown also mitigates the effect of a delay in the initial response. Compliance with lockdown must be at least 80-90%, however, or epidemic control will be lost.

**Conclusions:** A SARS-CoV-2 outbreak will spread rapidly in remote communities. Prompt case detection with quarantining of extended-household contacts and a 14-day lockdown for all other residents, combined with exit testing for all, is the most effective strategy for rapid containment. Compliance is crucial, underscoring the need for community supported, culturally sensitive responses.

## Background

The SARS-CoV-2 pandemic continues to cause significant morbidity and mortality worldwide, disproportionately affecting vulnerable and disadvantaged groups such as those of lower socio-economic status, or with comorbidities [1]. Protecting such groups must be a priority. As of mid-2020, Australia was in a favourable position until a resurgence of cases in Melbourne highlighted ongoing susceptibility to outbreaks [2]. A city-wide lockdown has followed, with strict lockdowns imposed on several crowded public housing complexes. No cases of community transmission have yet occurred in remote Australian Aboriginal and Torres Strait Islander communities.

Within Australia, Aboriginal and Torres Strait Islander peoples (hereafter respectfully referred to as ‘Aboriginal’) are significantly more vulnerable to severe COVID-19, due to a high prevalence of comorbidities associated with severe clinical outcomes [3]. The incidence of chronic respiratory diseases is 1.2 times higher than for non-Aboriginal Australians, type 2 diabetes 3.3 times higher, and chronic kidney disease 3.7 times higher [4]. SARS-CoV-2 transmission is likely to be more intense within remote communities due to crowded housing, larger family sizes, inadequate hygiene facilities, and residence across multiple dwellings (4-7). These communities are also further from specialist health services, with SARS-CoV-2 tests needing to be transported resulting in delays. Previous influenza outbreaks in these communities have underscored their vulnerability. During the 2009 H1N1 pandemic, hospital and ICU admissions for Aboriginal people were 12 and 5 times higher, respectively, than for non-Aboriginal Australians [5]. Similarly, First Nations Americans of the Navajo Nation have suffered the highest rates of SARS-CoV-2 infection in the USA, with case fatality rates more than triple that of Australia overall [6]. The consequences of overcrowding and disadvantage have been demonstrated in Singapore, where migrant workers in overcrowded dormitories suffered from infection rates of up to 20% [7].

In Australia, protection of remote Aboriginal communities was prioritised early, including establishment of strict movement controls in consultation with communities, within designated biosecurity zones [8]. A national advisory body, the Aboriginal and Torres Strait Islander Advisory Group on COVID-19 (the IAG), co-chaired by the Department of Health and the National Aboriginal Community Controlled Health Organisation, provides evidence-based and culturally safe guidance for COVID-19 preparedness and response to the government and other key stakeholders, with a view to locally led adaptation within each community [9]. This group liaises with peak national health advisory bodies on COVID-19 and commissioned the work that we present here to help inform optimal public health response strategies in remote settings.

## Methods

We compare plausibly implementable strategies in a remote Aboriginal community, examining the impact of alternative scenarios in an outbreak response, including: initial delays with testing; differing definitions of case-contacts and consequent quarantine strategies; community-wide lockdowns; and exit testing strategies.

### Participatory approach

A participatory approach was employed throughout this study. All of the SARS-CoV-2 outbreak response scenarios explored were designed through iterative engagement between the academic investigators, the IAG, and other public health end-users to ensure cultural sensitivity, and to maximise the relevance and uptake of findings.

### Population assumptions

An individual-based model, repurposed from a framework developed to examine dynamics of sexually transmitted infections in remote Australia, is used to explicitly represent each community member [10]. Community sizes comprising 100, 500, 1,000 or 3,500 people are modelled, with results presented here focusing on communities of 1,000 people but noting key differences.

We adopt the population household structure described by Chisholm et al.[11], whereby individuals have family connections across multiple dwellings. Each person’s time at home is distributed between a main dwelling (core) 66% of the time, second dwelling (regular) 23%, and third dwelling (on/off) 9%. The remaining 2% is spent at a randomly allocated dwelling. The frequency of contact, and therefore likelihood of transmission, is higher between individuals within the same dwellings. Section 3 (Appendix) provides a summary of household distribution and contact rates.

### Epidemic assumptions

The disease model follows a susceptible, infectious, recovered paradigm (see Figure 1). We assume infectiousness commences 48 hours prior to symptom onset on average [12] and ceases with symptom resolution. Table A-1 of the appendix summarises key transmission parameters. The basic reproduction number *R*_*0*_ was calibrated to centre around 5, based on similar contexts [13-15] and allowing for enhanced mixing anticipated in overcrowded households [16-18] (Section 2 of appendix). We conservatively assume that only half of infected patients will self-present for testing, due either to minimal/no symptoms, fear, or stigma.

**Figure 1:**
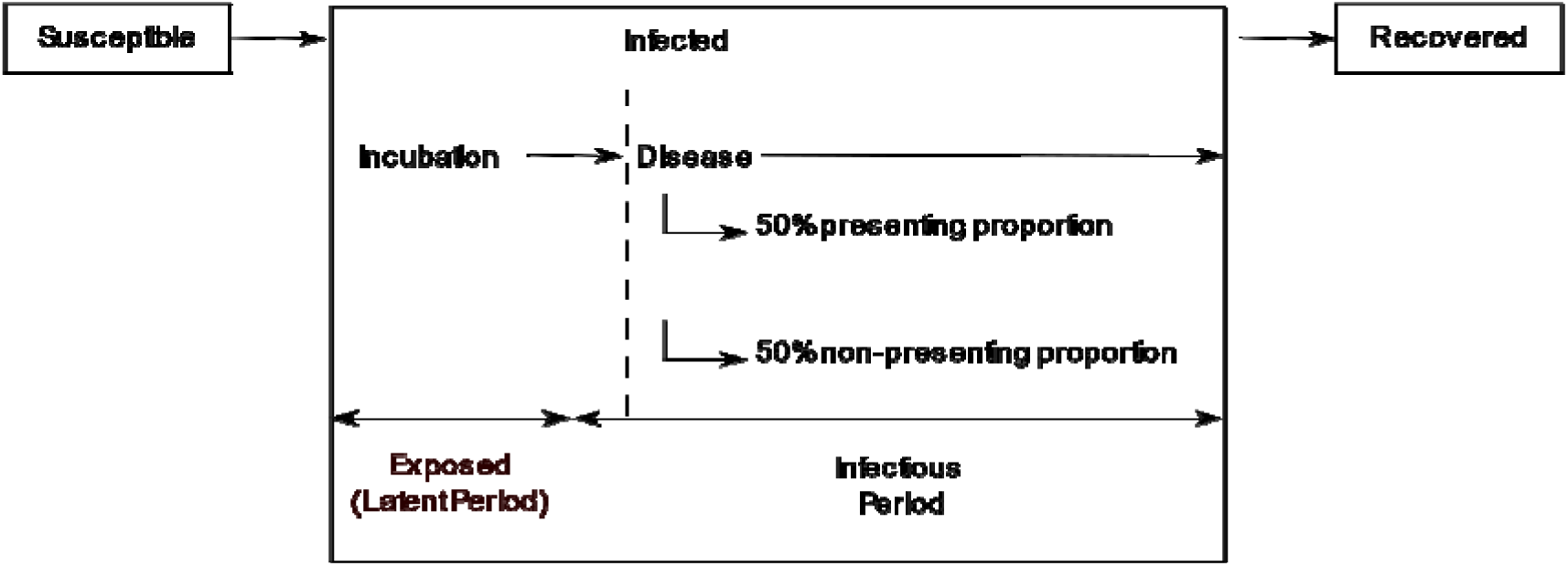
Susceptible, Infected, Recovered Model Assumptions. After exposure to SARS-CoV-2 a proportion of susceptible individuals become infected, entering the incubation phase before proceeding to the disease phase. 50% of individuals in the disease phase are assumed to spontaneously present to clinical services, the ‘presenting proportion’. The remaining ‘non-presenting proportion’ (those with few or no symptoms or who avoid health services due to fear or stigma) will only be identified through active case finding and testing efforts as part of the public health response. We assume infectiousness commences 48 hours prior to onset of symptoms (if they occur) and persists until resolution of symptoms. While we do not explicitly split out asymptomatics from the non-presenting proportion, we conservatively assume that they are as infectious as individuals with symptoms.

### Intervention assumptions

The impact of a multi-layered public health response is assessed following identification of the index case. *Cases* (those who test positive for SARS-CoV-2) are assumed to be isolated immediately and effectively. *Contacts* of cases, as variously defined below, are quarantined alone and assumed to be completely separated from others.

### 1. Contact definitions and quarantine

Two broad strategies for contact definition are assessed as per Figure 2. For household-based, we define *immediate* household contacts as those who share the same dwelling at the time of tracing; *extended* household contacts are those who share other dwellings a case may inhabit. For history-based contact tracing, contacts are those identified over the prior 2 days (close and casual).

**Figure 2:**
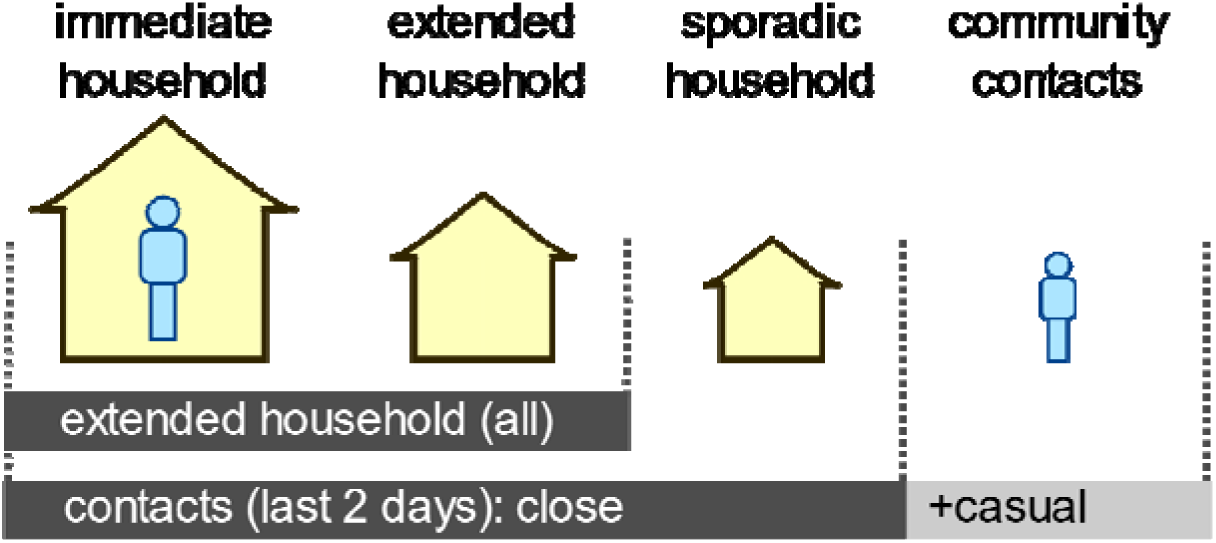
Definitions of contacts. Household-based contacts include the index case’s immediate and extended households defined by dwelling. History-based contact tracing relies on active contact tracing for the preceding 2 days, including household (close) contacts and community (casual) contacts.

### 2. Lockdown of community

Concurrent with the quarantining of contacts, the impact of a 14-day lockdown of all households within a community is modelled. Under lockdown, individuals remain in their core dwelling and can mix with other residents, but *not* with other households or the wider community. The effect of non-compliance is assessed.

### 3. Testing

Initial testing of individuals for SARS-CoV-2 occurs following clinical presentation, or after identification as a contact. We assume a 2-day delay between taking the test and the public health response being initiated due to logistical factors. We also assume 100% test sensitivity. The impact of subsequent testing is examined for various scenarios:

- *Entry* testing of all contacts when commencing quarantine
- *Clearance* testing prior to release from *quarantine* for all contacts (on day 12 of 14, assuming a 2-day delay)
- *Clearance* testing prior to release from *isolation* for all cases (on day 8 of 10, assuming a 2-day delay)
- *Clearance* testing prior to release from *lockdown*

Positive tests at any point are treated as new cases, triggering a further round of contact tracing with subsequent isolation and quarantine (+/-lockdown).

## Results

### Impact of delays to case finding

We assume a scenario in which an initial case enters the community while pre-symptomatic and is detected only on subsequent self-presentation and testing. The number of infected individuals likely present in the community by the time the first case is identified is summarised in Table 1. Projections for multiple initial cases being identified are also shown. Figure A-2 (Appendix) summarises projected numbers if a lower proportion of cases self-present to health services.

**Table 1:** Impact of delays to case finding. Scenarios are shown for a range of community sizes, summarising the number of currently infected individuals and the cumulative number of infected individuals present by the time that the initial one, two or five cases are identified. Medians with interquartile ranges (in brackets) are reported from 100 simulations.

| Population size | One case identified |  | Two cases identified |  | Five cases identified |  |
| --- | --- | --- | --- | --- | --- | --- |
|  | Current infected individuals | Cumulative infected individuals | Current infected individuals | Cumulative infected individuals | Current infected individuals | Cumulative infected individuals |
| 100 | 9 (5, 16) | 32 (15, 47) | 18 (11, 25) | 52 (38, 65) | 37 (28, 46) | 84 (71, 90) |
| 500 | 7 (2, 15) | 29 (9, 55) | 20 (12, 30) | 73 (50, 104) | 48 (37, 63) | 162 (134, 207) |
| 1000 | 6 (3, 14) | 27 (10, 59) | 19 (11, 27) | 72 (46, 100) | 50 (33, 68) | 184 (131, 235) |
| 3500 | 7 (4, 11) | 22 (9, 42) | 18 (11, 25) | 66 (44, 105) | 49 (35, 67) | 187 (144, 247) |

#### Impact of delays to case finding

### Impact of definition of contacts, and quarantine strategies

In the absence of entry and clearance testing, the extended household-based contact tracing and quarantine strategy results in a peak infection prevalence of approximately 40%, versus 50% for the history-based quarantine strategy (Figure 3, upper panels). The addition of *entry* testing to quarantine reduces the peak infection prevalence for the extended household-based strategy to approximately 10%, versus 40% for the history-based strategy (middle panels). Adding both *entry and clearance* testing results in a small additional benefit to the extended household strategy (largely in the reduction of outbreak duration), but no significant benefit to the history-based strategy.

**Figure 3:**
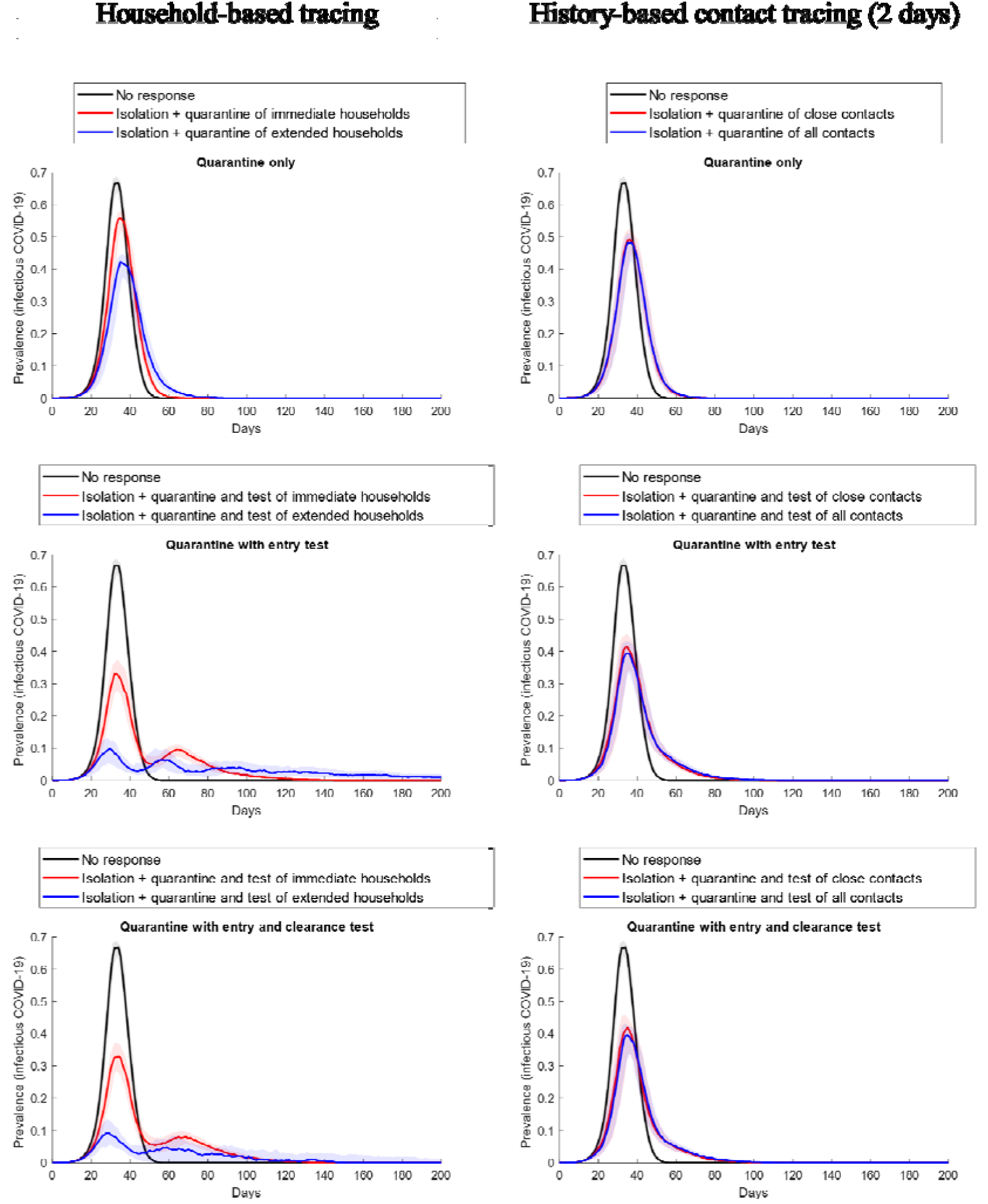
Impact of different contact tracing strategies: epidemic curves for a community of 1000 people, comparing the household-based tracing strategy, at left, with the history-based tracing strategy (for the prior 2 days) at right. Lines represent the median value and shaded areas the interquartile ranges from 100 simulations.

The impact of clearance testing with various quarantine strategies on total infection numbers (i.e. not just peak prevalence) is greatest for the extended household approach (Table 2). In all other strategies, more than 90% of the community are ultimately infected, with or without testing. For extended household quarantine *without* clearance testing, 83% are infected, ∼87,000 person-days spent in quarantine and >4000 tests performed. The addition of clearance testing results in ∼66% being infected, fewer person-days in quarantine (∼51,000) but more tests (13,551), making it the most effective strategy.

**Table 2:** Impact of clearance testing on contact tracing and quarantine strategies for a community of 1,000 people. Size of outbreak (total cumulative infections), quarantine person-days (per 1,000 population), and total tests performed during outbreak are shown. Medians are reported, with interquartile ranges (in brackets) from 100 simulations. Note that ‘infections’ refers to all individuals with SARS-CoV-2 infection (whether tested and known to health services or not), whereas ‘cases’ refers to those with SARS-CoV-2 infection who have tested positive, i.e. have been identified.

| Strategy | No clearance testing |  |  | Clearance testing undertaken |  |  |
| --- | --- | --- | --- | --- | --- | --- |
|  | Total cumulative infections | Quarantine person-days (first year) | Total tests performed during outbreak (first year) | Total cumulative infections | Quarantine person-days (first year) | Total tests performed during outbreak (first year) |
| No response | 999 (999-999) | N/A | 447.0 (435.5, 458.0) | 999 | N/A | N/A |
| Quarantine of immediate household contacts (with case isolation) | 922.0 (907.5, 936.5) | 29595.5 (28101.5 - 31175.0) | 1957.5 (1867 - 2027) | 922.5 (905.0, 933.0) | 22500.5 (21469.0 - 23306.0) | 7526.0 (7336 - 7743) |
| Quarantine of extended i.e. all household contacts (with case isolation) | 831.5 (751.0, 871.0) | 86825.0 (70334.5 - 97662.5) | 4042.5 (3463 - 4305) | 655.0 (267.5, 821.0) | 50958.0 (13511.5, 67786.0) | 13551.5 (4929.5, 16729.5) |
| Quarantine<br>of *close*<br>contacts<br>based on<br>history (past<br>2 days) | 937.0 (929.0,<br>945.0) | 10776.5<br>(9551.5 -<br>11564.5) | 1530.5 (1441<br>- 1586) | 930.5 (917.0,<br>939.5) | 9445.5<br>(8541.5,<br>10191.5) | 4673.5<br>(4549.5,<br>4780.5) |
| Quarantine<br>of *all*<br>contacts<br>based on<br>history (past<br>2 days) | 930.0 (917.0,<br>941.0) | 11887.0<br>(11180.0 -<br>12831.5) | 1614.5 (1550<br>- 1667) | 919.0 (904.5,<br>931.5) | 10662.0<br>(9718.0,<br>11768.5) | 4842.5<br>(4741.0,<br>4957.0) |

#### Impact of clearance testing on contact tracing and quarantine strategies

### Impact of community lockdown

Building on the extended-household quarantine strategy, the impact of *lockdown on all remaining households* (i.e. non-quarantined households) is shown to reduce both epidemic peak and duration – particularly if clearance testing is undertaken (Figure 4). Clearance testing from quarantine and lockdown is the most effective strategy to avert subsequent waves of infection in the community (green line). Entry testing is assumed for all these scenarios.

**Figure 4:**
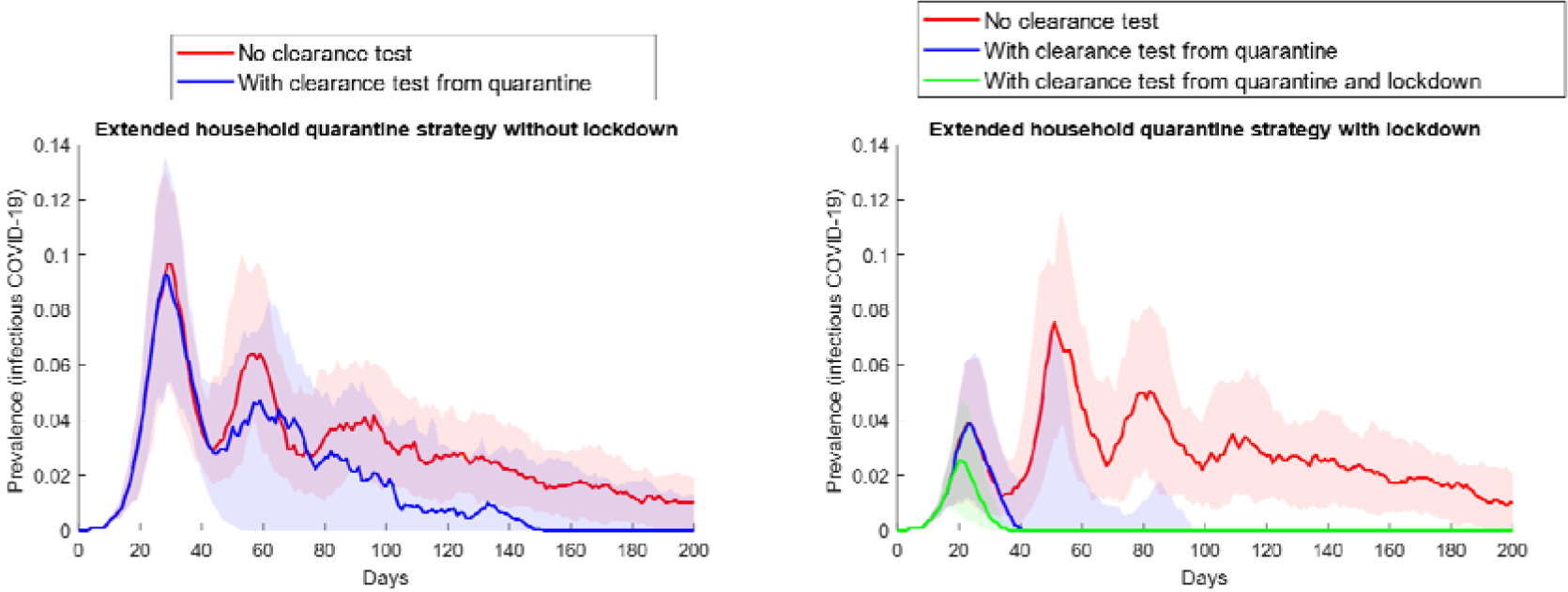
Impact of community lockdown on the extended household-based quarantine strategy, for a community of 1,000 people. The panel at left shows epidemic curves for the non-lockdown scenario (a composite of outputs from Figure 3), with and without clearance tests from quarantine. The panel at right shows these scenarios with lockdown. Entry testing is assumed for all individuals. Lines represent the median value and shaded areas the interquartile ranges from 100 simulations.

Lockdown with clearance testing is also the most effective strategy to reduce *total* cumulative infections, when applied alongside the extended household quarantine strategy with clearance testing (Table 3). Without any clearance testing (top row), lockdown alone has little impact on total infections (>800), quarantine person-days (>85,000), or tests (∼4,000). Adding clearance testing to quarantine only (middle row) results in fewer infections with lockdown added (89 versus 655), similar quarantine person-days (∼5,000), and far fewer tests (1,402 versus 13,551). Undertaking clearance testing for both lockdown and quarantine (bottom row) results in only 35 infections in total, fewer quarantine person-days, and ∼2,500 tests – the optimal strategy.

**Table 3:**
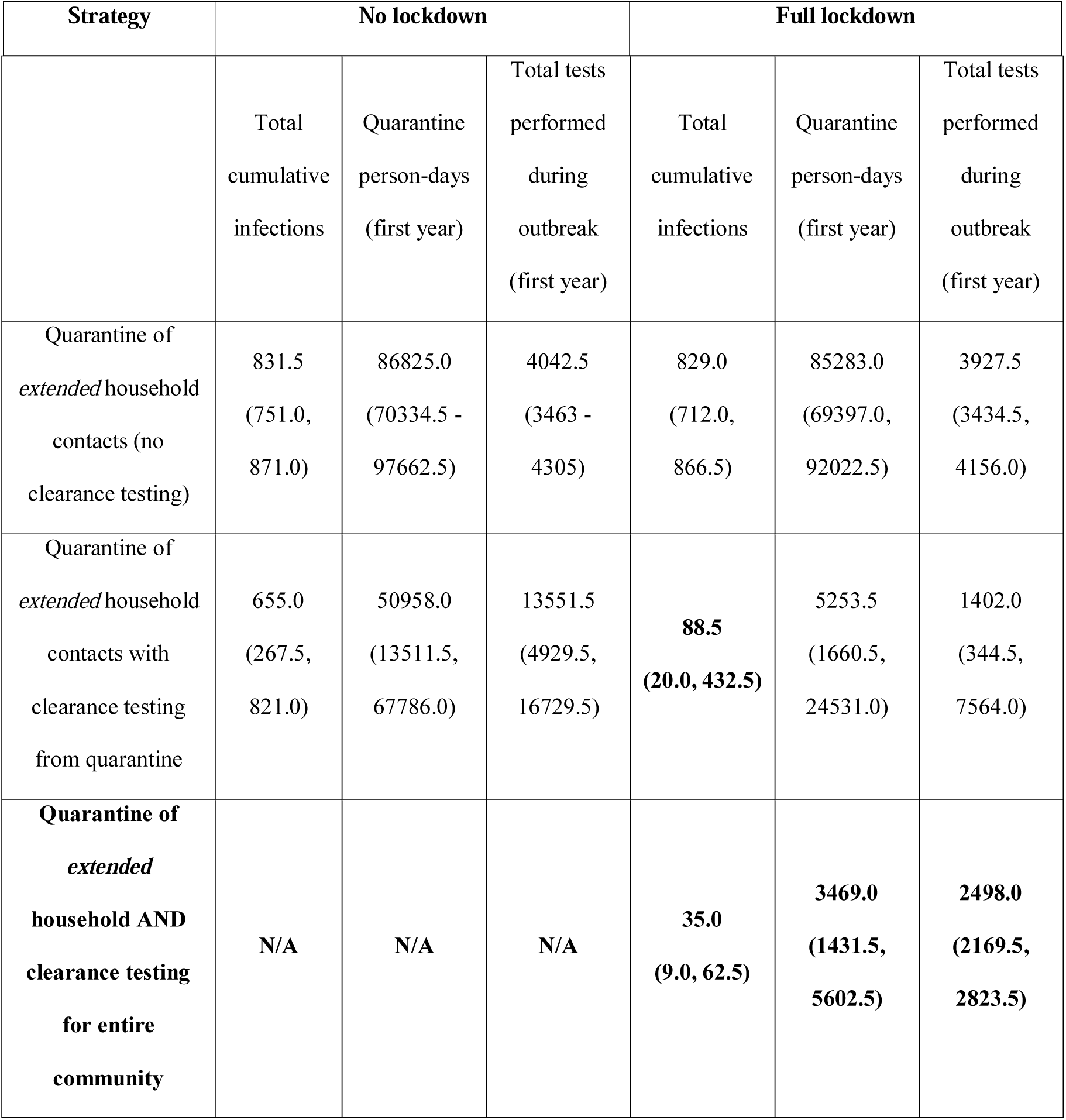
Impact of lockdown and extended household quarantine, combined with various testing strategies, for a community of 1,000 people. The effect on size of outbreak (total cumulative infections), quarantine person-days (per 1,000 population), and total tests performed during outbreak are shown. Figures are medians, with interquartile ranges.

#### Impact of lockdown and extended household quarantine combined with various testing strategies

### Impact of lockdown on delays with intervention

The effect of delays between the identification of cases and implementation of interventions is mitigated by the addition of lockdown (Figure 5). For the extended household quarantine scenario, increasing the delay from 2 to 4 days in the absence of a lockdown, causes infection prevalence to increase from <10% to ∼25%, and to ∼45% with a 6-day delay (left panel). The addition of lockdown results in a peak prevalence of <15%, even with a 6-day delay to implementation (right panel).

**Figure 5:**
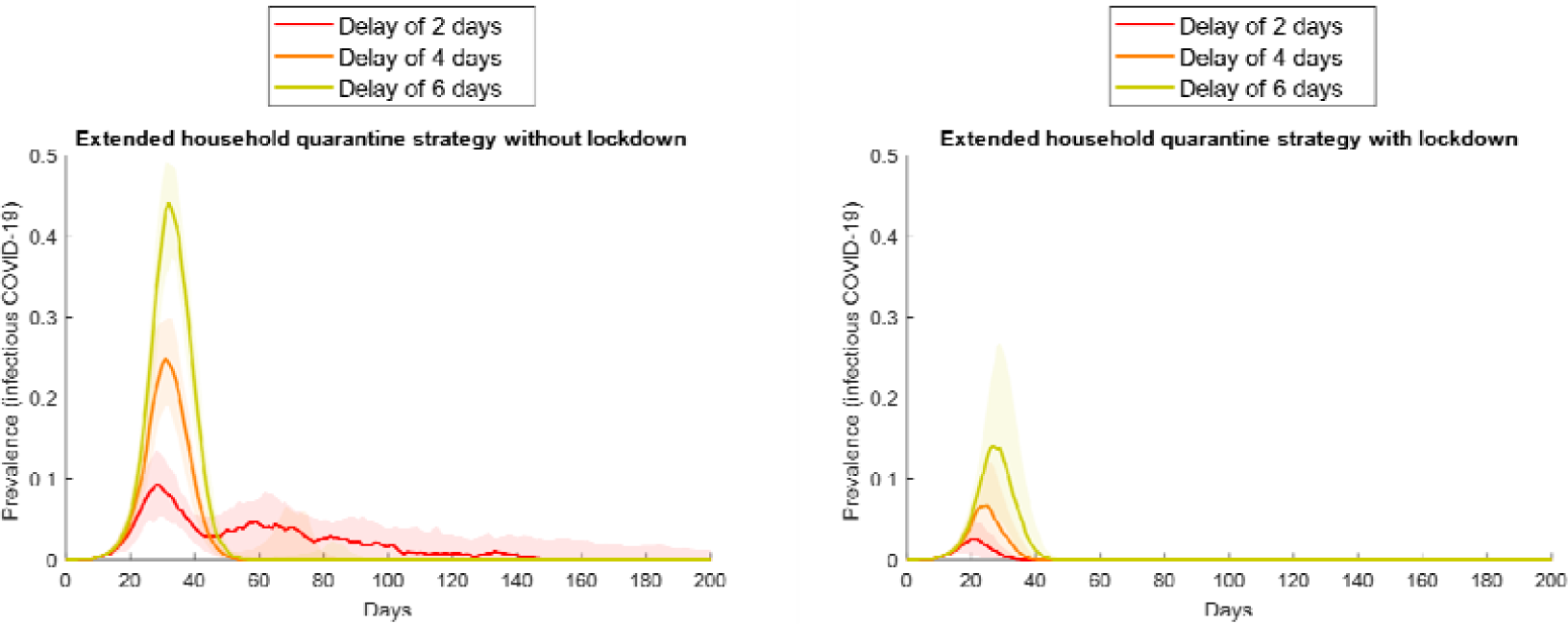
Impact of lockdown on outbreak control, comparing delays in the response following testing of index case. Epidemic curves shown for the extended household quarantine scenario in a community of 1,000 people, with entry and clearance testing. Initial outbreak response following the identification of the index case is delayed by 2, 4 or 6 days; the no-lockdown scenario is shown at left, with lockdown at right. Median values and interquartile ranges (shaded) from 100 simulations are shown.

### Impact of compliance with lockdown

Loss of epidemic control occurs even in the optimal strategy (lockdown alongside the extended household quarantine strategy, with entry and clearance testing) when compliance for individuals with lockdown falls below 80% (Figure 6).

**Figure 6:**
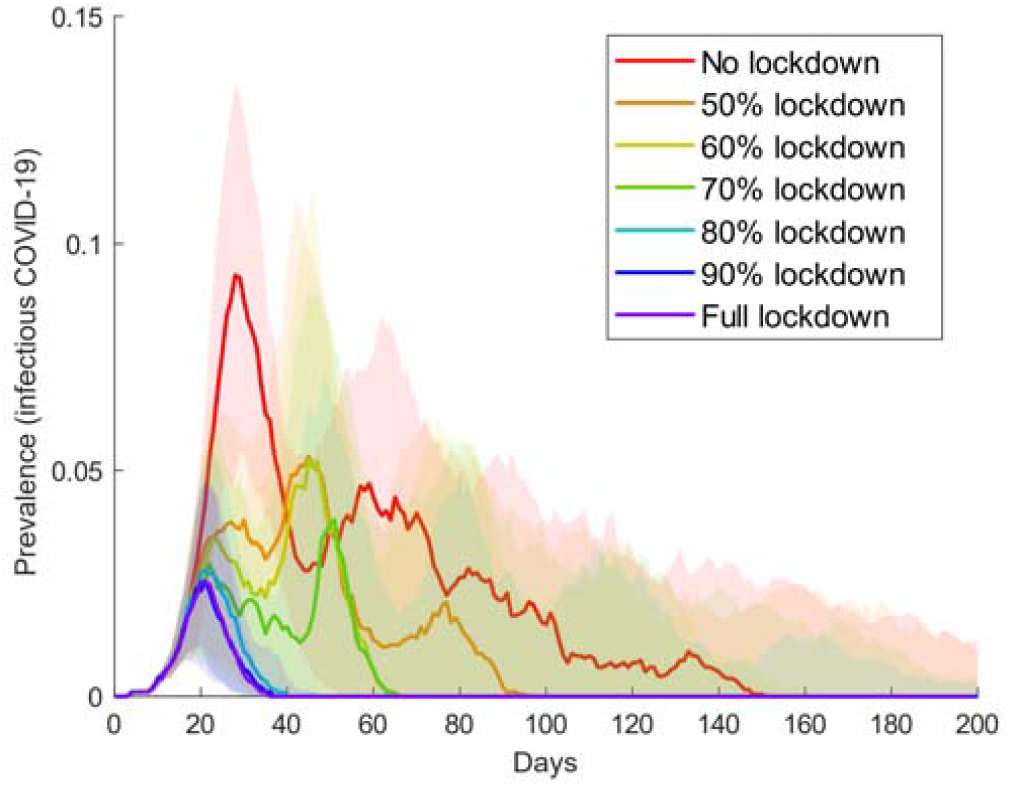
Impact of compliance with lockdown on a community of 1,000 people. Epidemic curves for the extended household quarantine strategy (with entry and clearance testing), with various levels of individual compliance with community lockdown. Median values and interquartile ranges (shaded areas) from 100 simulations shown.

### Impact of community size on the effect of lockdown

For small communities of 100, lockdown has little additional impact as most are already quarantined due to extended household membership (Figure 7). For communities of 500, lockdown reduces peak prevalence from ∼10% under the extended household quarantine strategy to ∼5%. Greatest benefit is seen in very large communities (3,500), where peak prevalence is reduced from ∼10% to less than 1%, and subsequent waves of infection are suppressed.

**Figure 7:**
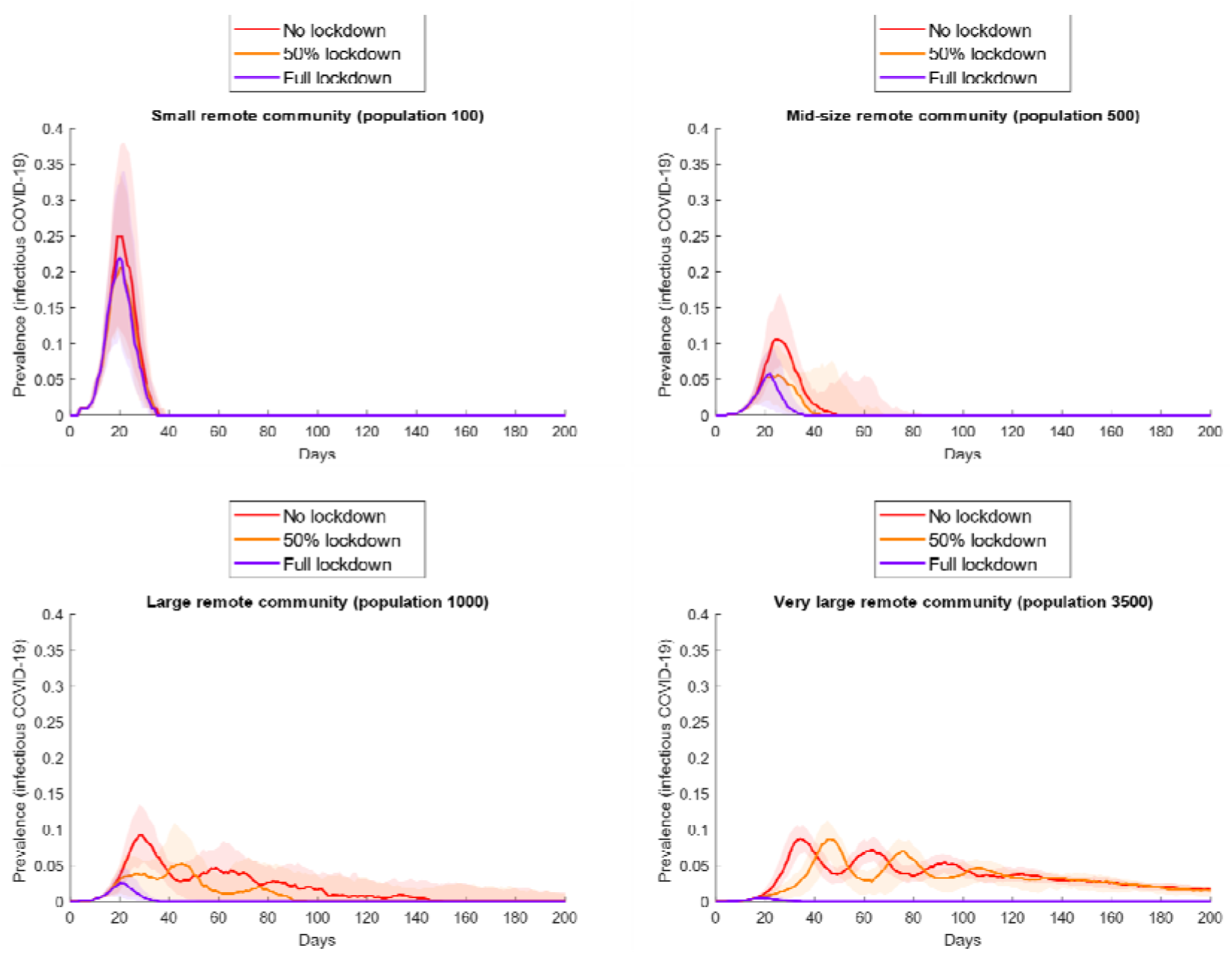
Impact of lockdown on communities of varying size. Epidemic curves for the extended household quarantine scenario (with entry and clearance testing), with perfect lockdown; lockdown with 50% compliance; and no lockdown. Median values (lines) and interquartile ranges (shaded areas) from 100 simulations shown.

## Discussion

Prompt case finding is essential to prevent a SARS-CoV-2 outbreak in a remote Aboriginal community. A high transmission propensity, due to interconnected and often crowded households, means that in an unmitigated scenario the majority of the community would be rapidly infected. By the time early cases are identified, active infections in the community may be up to ten-fold higher. We assume only half of all infected patients will self-present to health services for testing, due to absent or minimal symptoms, fear, or stigma. This may be an overestimate, but evidence that pre-symptomatic transmission may contribute >40% of SARS-CoV-2 transmission exists [12, 19]. This non-presenting proportion may not be detected using a passive case finding approach, although a high prevalence of other co-morbidities may result in non-COVID related presentations resulting in ‘co-incidental’ case detection. Higher non-presenting proportions would lead to poorer mitigation in all scenarios, and vice versa (see Appendix).

Of the contact tracing strategies, quarantining *extended household* members (residents of all dwellings used by the case) is the most effective strategy for constraining the initial outbreak, reducing peak prevalence from 60-70% to ∼10% (Figure 3). However, large numbers of people must be quarantined for extended periods and infections may resurge when community mixing resumes, with overall community attack rates exceeding 80% (Table 3). Clearance testing modestly reduces this attack rate to 65%. Lockdown of all non-quarantined households for 14 days, concurrent with this quarantine strategy, results in the greatest likelihood of definitive outbreak control. Peak prevalence of the initial outbreak is less than 5%, and the overall attack rate less than 10%. Clearance testing from lockdown further improves control, preventing subsequent waves of infection due to undetected infections being released (Figure 4): overall infections are constrained to <5% with clearance testing, versus >80% without. This strategy also requires fewer tests due to prompt suppression, fewer person-days in quarantine, and remains effective with delays of up to 6 days (Figure 5). Larger communities benefit most from lockdown, with the effect dampened in smaller communities (100-500) by the large proportion already in quarantine. Compliance with lockdown must be at least 80-90%, or epidemic control will be lost.

Our findings are consistent with recent guidelines for a ‘contain and test’ strategy developed by Central Australian health organisations [8], which acknowledge that symptom-based case identification will be insufficient, and endorse active case finding and lockdown with multiple rounds of voluntary testing. Analyses of SARS-CoV-2 outbreaks overseas also support the effectiveness of lockdowns. In the Italian town of Vo, researchers concluded that a 14-day lockdown reduced transmissibility of infections (including asymptomatic) by 82-98% [20]. Lockdowns in Wuhan contributed to a significant decrease in spread [21], and an analysis of French data suggested that over 80% of potential COVID-19 deaths were averted by their lockdowns [22]. Recent modelling from the UK, examining the impact of delays with testing and contact tracing, suggests that if cumulative delays exceed 3 days for these processes, control of an outbreak is unlikely [23].

The participatory process employed between this study’s investigators, the IAG, and other public health end users throughout, have allowed for direct feedback of our findings and incorporation into IAG guidelines [9], and collaborative development of plain-language messaging for health providers and community members. Prompt case finding and a rapid public health response will be critical for effective control, with access to decentralised point-of-care testing (e.g. *GeneXpert*) facilitating this. Local planning and preparation should occur in advance, and must involve community members to ensure cultural appropriateness, local support and community control. Early patient presentation should be encouraged, and testing, contact tracing and isolation/quarantine guidelines and facilities clarified. The extensive public health response required to achieve best outcomes necessitates prior preparedness planning to ensure that the significant logistical and human resources support needed can be rapidly mobilised. Throughout an outbreak, community trust must be preserved in order to maximise compliance; in particular, the historical context and consequent sensitivities regarding enforced lockdowns in remote Aboriginal communities must be kept foremost in mind in the design and implementation of such strategies.

### Limitations

Our model is informed by simplifying assumptions derived from observational data regarding population structure and mixing. Other ‘real world’ mixing opportunities (e.g. schools and workplaces) have not been explicitly included. Assumptions regarding transmission dynamics are derived from non-Aboriginal populations, but where possible we have erred on the side of caution. The high *R*_*0*_ to which the model is calibrated is based on early estimates from Wuhan and amplified to reflect the propensity for intense transmission in remote households. We assume perfect sensitivity and specificity of testing throughout the infectious period. Morbidity and mortality outcomes have not been estimated in this model, or the anticipated demand on health resources (testing requirements aside).

We assume that cases in isolation and contacts in quarantine will have no contact with others (i.e. will not transmit SARS-CoV-2). This may not be possible to achieve, but by representing this ideal we assess the maximum effectiveness of these measures and demonstrate the added value of lockdown.

## Conclusions

Remote Australian Aboriginal and Torres Strait Islander communities have the potential to be severely impacted by COVID-19, due to factors favouring increased transmission and disease severity. Our modelling affirms the need for early case detection, as multiple secondary infections are likely already presents by the time an index case is identified. Quarantining of extended household contacts, together with 14-day community-wide lockdown with clearance testing, are the most effective strategies in limiting the outbreak.

## Data Availability

The modelling code is publicly available at Zendo/GitHub at the following DOI: 
https://doi.org/10.5281/zenodo.4057288

https://doi.org/10.5281/zenodo.4057288

## Abbreviations

COVID-19: Coronavirus Disease 2019
H1N1: Influenza A virus subtype H1N1
IAG: Aboriginal and Torres Strait Islander Advisory Group on COVID-19
ICU: Intensive care units
SARS-CoV-2: Severe acute respiratory syndrome coronavirus 2

## Declarations

### Ethics approval and consent to participate

Not applicable

### Consent for publication

Not applicable

### Availability of data and materials

Project name: Package_RMP

Project home page: https://github.com/The-Kirby-Institute

Operating system(s): Platform independent

Programming language: Java

### Authors’ contributions

All authors contributed to the design of the study and interpretation of results. BBH implemented and analysed the model. DB and BBH drafted the manuscript. JMcV and DGR initiated the study. All authors read and approved the final manuscript.

## Acknowledgements

Not applicable

