## Appendix for "Modelling testing and response strategies for COVID-19 outbreaks in remote Australian Aboriginal communities"

### Appendix: COVID-19 Remote Model

#### 1. Summary

We developed an individual-based model to simulate SARS-CoV-2 transmission in a setting representing remote Aboriginal communities in Australia. We consider communities of size 100, 500, 1000 and 3500 individuals, each representing small, median, large, and very large remote communities, respectively.

The population of each community comprises individuals of both genders aged 0-80 years, with the age and infection status of each individual tracked and updated daily. The population distribution for each age and gender group is maintained throughout the simulation, with aged-out individuals (those who would attain the age of 80 years) replaced by a new individual of the same gender and of age zero. The gender-age distribution is based on Australian Bureau of Statistics data for Aboriginal and Torres Strait Islander people living in remote regions in 2019 [1].

Transmission of infection can occur if there is contact between an infectious individual and a susceptible individual. See Section 2, “Infection (SARS-CoV-2)” for a detailed description of how SARS-CoV-2 transmission is defined in this model.

Individuals residing in the same community are grouped into households where person-to-person contact is more frequent than in the general community, thereby providing more opportunity for successful transmissions. See Section 3, “Household” for a detailed description of how contacts are defined in this model.

Individuals can be tested for SARS-CoV-2 infection if they seek testing themselves (because of symptoms), through contact tracing (if they are a contact of a case), or through clearance testing from isolation, quarantine or lockdown. See Section 4, “Contact tracing”, for a description of the types of contact tracing defined in this study, Section 5 “Testing” for the different types of testing defined, and Section 6, “Response”, for the responses enacted when an individual receives a positive test result.

The model is programmed in Java and the full code is available on GitHub at the following address <https://github.com/The-Kirby-Institute>.

#### 2. Infection (SARS-CoV-2)

The natural history of SARS-CoV-2 is captured in the model as illustrated schematically in Figure A‑1. Susceptible individuals who become infected following contact with an infectious individual initially enter a latent stage during which they are not yet infectious. They then enter the infectious stage, during which time they can infect others through contact. Infectious individuals then proceed to a post-infectious stage where they remain infected but are no longer infectious, before proceeding to recovery. We assume that recovered individuals are no longer susceptible to infection, i.e., they have acquired immunity to re-infection.

At the time of writing, the length of the incubation period (i.e., time for symptoms to appear after infection/exposure) and the length of the latent period (i.e., time for an infected/exposed individual to become infectious) is largely uncertain, therefore in this model the onset of symptoms is not considered explicitly as an infection stage, but as a separate condition that determines how likely it is that an infected individual will seek testing. We assume infected individuals will only seek testing if they have symptoms and that they are no longer infectious once symptoms resolve.

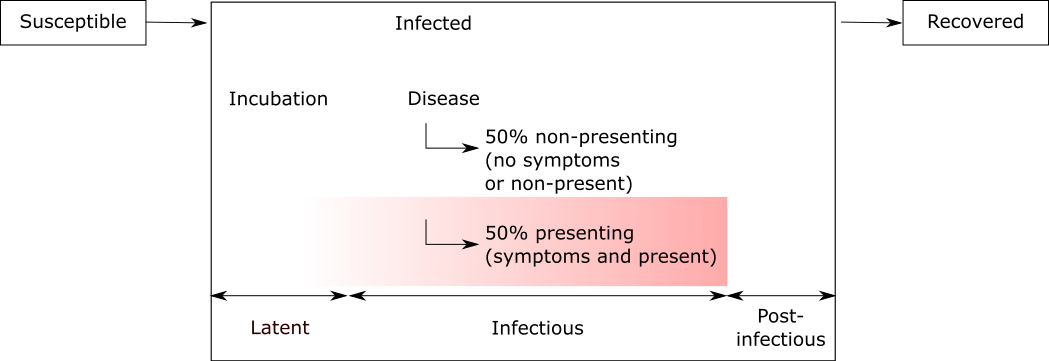

Figure A‑1: Schematic diagram illustrating the natural history of SARS-CoV-2 infection as captured in the model. Individuals can transmit SARS-CoV-2 during the infectious period. Some infected individuals develop symptoms following incubation and seek a test (red region in the figure). Note that the onset of symptoms may occur before or after an individual becomes infectious, but on average individuals become infectious 48 hours prior to onset of symptoms.

The infection parameters used in the model are assigned on a per-infection basis and are sampled from distributions as listed in Table A‑1.

Table A‑1: Parameters related to SARS-CoV-2 infection

| **Parameter** | **Distribution sample from** | **Source** |
| --- | --- | --- |
| Latent period (days) | Uniform distribution range 3 to 6 | [2-5], expert opinions. |
| Incubation period (days) | Weibull distribution with mean of 6.4 and SD of 2.3 |  |
| Infectious period (days) | Weibull distribution with mean of 10 and SD of 4 |  |
| Post-infectious period (days) | Uniform distribution range 0 to 10 |  |
| Immunity | Infinity (i.e. no re-infection occurs post infectious period | Assumption |
| Transmission probability per contact | Uniform distribution range 0.15 to 0.20 | Calibrated such that a R_0_ of 5.0 is maintained across the population. See the section on calibration for full explanation |

We assume that a substantial proportion of infected individuals (50%) will not seek a test even if they have symptoms due to either mildness of symptoms, difficulty in accessing healthcare in remote settings, and/or fear or stigma associated with testing. Therefore, instead of classifying infected individuals as either symptomatic or asymptomatic, we classify infected individuals as either “test seeking” or “non-test seeking”, with the latter including those who have symptoms but do not seek a test. The proportion of infected individuals who seek a test is assumed to be 50% in our baseline analysis, but in “Supplementary results” presented at the end of this appendix, we examine outcomes when this proportion is varied.

##### Model calibration

Model calibration is performed by adjusting the per-contact transmission probability. At the time of writing, little is known about the per-contact transmission probability for SARS-CoV-2, which may vary by infection stage and severity of symptoms. In the current model we make the parsimonious assumption of a uniform per-contact transmission probability for the entire infectious period and note that this is likely a simplification.

The magnitude of the per-contact transmission probability is also uncertain. Early studies from Wuhan, China (where SARS-CoV-2 was first identified) suggested a basic reproduction number (R_0_) in the range 1.4 – 5.7 [6-8]. WHO estimated R_0_ to be in the range 1.4 – 2.5 in early January 2020, while a more recent review suggested R_0_ to be 3.3 across a range of modelling and non-modelling studies, with R_0_ lies between 2.0 - 3.0 among modelling studies of similar vein (i.e. studies that uses stochastic simulations) [9].

Furthermore, R_0_ is likely to be higher in remote Aboriginal communities in Australia compared to in communities in China due to their larger average household sizes – 7.7 co-inhabitants per household on average in remote communities [10] compared with 3.1 in China [11]. Based on this difference in average household size, we assume that the R_0_ for remote communities is 2 to 3 times higher than that estimated for Wuhan, and we calibrate the model (by adjusting the per-contact transmission probability) to produce an R_0_ of approximately 5.

#### 3. Household

At the beginning of each simulation, individuals in a given community are grouped in terms of households. The household structure is maintained for the entire simulation unless a household member is removed (e.g., due to aging out), in which case the removed member is replaced with a new individual of the same gender aged 0 years.

We have adopted the household structure described by Chisholm *et al*.[12], whereby individuals inhabit one of their pre-allocated dwellings on a given day or, less frequently, a randomly selected dwelling within the population. Each individual is considered to have close family connections across a total of three dwellings: their main dwelling (core), which they occupy 66% of the time; a second dwelling (regular), which they occupy 23% of the time; and third dwelling (on/off), which they occupy 9% of the time. Their remaining time in a household (i.e., 2%) is spent at a randomly allocated dwelling at the start of each time step. On a given day, an individual’s immediate household is defined as those individuals who inhabit the same dwelling as said individual on that day. An individual’s extended household is defined as those individuals who are members of said individual’s immediate household or are members of their core, regular or on/off dwellings.

The size of a core household (the number of individuals who are assigned the same core dwelling) is based on the study by Bailie and Wayte, which estimated the mean household size for a remote community to be 7.7.[10] The number of households and household size for each community is then determined by sampling from a Poisson distribution. The output snippet below shows some summary statistics (mean, interquartile range, minimum and maximum) on core household size for a typical simulation:

```
Pop size: [10000, 100, 500, 1000, 3500]
Household:
For Loc #1
# household = 14
Size of households = 8.143 [7.000, 9.250] (Min, Max = 4, 10)
For Loc #2
# household = 70
Size of households = 8.143 [6.000, 10.000] (Min, Max = 2, 13)
For Loc #3
# household = 139
Size of households = 8.194 [7.000, 10.000] (Min, Max = 2, 16)
For Loc #4
# household = 441
Size of households = 8.937 [7.000, 11.000] (Min, Max = 3, 18)
```

By default, individuals have daily contact with members of their immediate household and can have additional contact with individuals outside of their immediate household (randomly selected from a pool of individuals who are not immediate household members). The non-immediate household contact rate is based on the frequencies of face-to-face contact with family or friends living outside the household as reported by the Australian Bureau of Statistics and is summarised in Table A‑2 [13].

Table A‑2: The frequency of non-immediate household contact (represented in terms of number of contacts per day) and the proportion of regional and remote residents within each category. For example, a rate between 1/30 and 1/7 means that group of individuals will have at least 1 non-household contact per 30 days to 1 non-household contact per 7 days.

| Frequency of non-household contact per day | Regional (%) | Remote (%) |
| --- | --- | --- |
| 1 | 20.7 | 25.0 |
| 1/7 to 1 | 54.5 | 50.6 |
| 1/30 to 1/7 | 15.9 | 16.2 |
| 1/90 to 1/30 | 5.2 | 4.2 |
| 0 | 3.7 | 4.1 |

Infection can be transmitted as a result of any contact. Refer to Section 2, “Infection (SARS-CoV-2)”, for a description on how SARS-CoV-2 transmission is defined in this model.

#### 4. Contact tracing

We refer to the individual with a positive diagnosis as a case. Contacts of a case are potentially identified through contact tracing/testing, and/or quarantined in the control scenarios we examine. For this study we define two forms of contact tracing: household-based, whereby contacts are identified and traced/tested on the basis of the case’s household membership; and history-based, whereby all of the case’s contacts in the last 2 days are traced/tested. The definition of each type of contact is summarised in Table A‑3.

In our baseline analysis, we assume a delay of 2 days between testing and initiation of the public health response (see Section 5, “Testing”).

Table A‑3: Individuals to be included in contact tracing under different contact tracing regimens

| Household-based | Immediate household | Extended household |
| --- | --- | --- |
|  | Individuals who are members of the case’s immediate household (i.e. living in the same house) at the time contact tracing is conducted | Individuals who are members of the case’s immediate household or are members of their core, regular or on/off households. |
| History-based | **Close contact** | **All contact** |
|  | Individuals who inhabited the same house (or houses) as the case in the last 2 days (measured as prior to the time contact tracing is conducted). | Individuals who are close contacts of the case or have had community contact with the case in the last 2 days (measured as prior to the time contact tracing is conducted). |

#### 5. Testing

Individuals can be tested in the following situations:

1. Spontaneous presentation for testing, for example due to onset of symptoms;
2. Testing to enhance case ascertainment among individuals identified as a contact of a case (i.e., someone who has a positive diagnosis);
3. Clearance testing, carried out to confirm infection free status prior to release from isolation, quarantine, or lockdown.

A case is identified if there is a positive diagnosis of SARS-CoV-2 infection. We assume tests are ideal (i.e. 100% sensitive) and will return positive diagnosis if the tested individuals are infected, even if they are not-yet infectious, have few or no symptoms, or are in the post-infectious phase.

Initially, we assume there is a time delay of 2 days between swabbing, access to the result, and the implementation of a response (e.g., isolation, contact tracing, population lockdown). In reality, different responses may be associated with longer delays. Therefore, we consider additional scenarios where the delay is 4 and 6 days.

##### Self-sought testing

We assume that an individual who seeks testing will only do so after the incubation period, on average 6.4 days after exposure. We also assume those who seek a test will do so before symptoms subside. Using the average duration of latent and incubation period from Table A‑1 above, we estimate symptoms will last approximately 8 days. We assume that individuals who spontaneously present for testing (half of all those infected) do so at a rate such that 99% will be tested within 8 days following the onset of symptoms; this distribution is weighted towards the early phases of infection such that 68% present within 2 days.

##### Contact tracing and testing

In some scenarios, contacts of an individual can be traced and tested if that individual is identified as a case (i.e. confirmed positive diagnosis). Refer to Section 4, “Contact tracing”, for the definition of contacts.

##### Clearance testing

For individuals who are already subject to contact restrictions (i.e., isolation, quarantine, lockdown), a clearance test may be conducted to confirm presence or absence of infection by the end of the restriction period. We assume isolation lasts 10 days, while quarantine and lockdown last 14 days. Therefore, to account for a diagnostic delay of 2 days, clearance testing is conducted on the 8^th^ day of isolation, and on the 12^th^ day for quarantine or lockdown.

#### 6. Response

Depending on the scenario being evaluated, one or more of the following responses may be triggered following the identification of a case (i.e., a positive diagnosis of SARS-CoV-2 infection):

- The case is placed in isolation for 10 days, with a clearance test, if applied, scheduled to occur on the 8^th^ day of isolation.
- All or a subset of contacts of the case (refer to Table A‑3 above for definition of contacts) are placed in quarantine for 14 days under different ascertainment strategies. A clearance test, if applied, is scheduled to occur on the 12^th^ day of quarantine.
- All or a subset of contacts of a case (depending on ascertainment strategy) are tested on entry to quarantine. Contacts returning a positive SARS-CoV-2 diagnosis then become new cases, triggering a new round of responses.
- Population lockdown (see below)

We assume any non-clearance test returning a positive diagnosis triggers further contact tracing, and hence multiple rounds of contact tracing and testing can occur from a single initial positive diagnosis (a case). In contrast, an individual returning a positive clearance test will remain in isolation for another 10 days until a subsequent clearance test (scheduled on 8^th^ day of the new isolation period) returns a negative result, but contact tracing will not be triggered as it will already have been conducted at the time the individual enters isolation/quarantine.

##### Lockdown

Population lockdown involves the quarantine of all members of the community not already in isolation/quarantine in their core households for a period of 14 days. Individuals in lockdown can mix freely with other members of their core household but not with other members of the community. In the scenarios we evaluate in this study, lockdown is triggered after the first confirmed case of SARS-CoV-2 infection in the population. Clearance testing, when applied, occurs on the 12^th^ day of lockdown and any individuals returning a positive diagnosis are subject to a further 10 days of isolation.

##### Contact restriction under isolation, quarantine, and lockdown

An individual in isolation, quarantine or lockdown will stay in their core dwelling for the entire restriction period, and will have limited contact with other individuals, hence preventing (in the case of isolation/quarantine) or reducing (in the case of lockdown) further SARS-CoV-2 transmission in the population. Contact restrictions are summarised in Table A‑4.

Table A‑4: Contact options for individual under various restrictions

| Nature of contact restriction | Reason for restriction | Can have contact with individuals cohabiting the immediate household at time *t* | Can have contact with  individuals outside the immediate household |
| --- | --- | --- | --- |
| No restrictions | Default | Y | Y |
| Isolation | Confirmed case – i.e. as a response to a positive diagnosis | N | N |
| Quarantine | Contact of a confirmed case | N | N |
| Lockdown | Population lockdown activated | Y | N |

#### 7. Supplementary results

##### Identification of first case

Timely identification of the first positive case is important as control strategies can only commence when there is confirmation of infection in the community. Furthermore, as the first positive diagnosis will be for an individual patient who has sought a test (due to onset of symptoms), the lag time for a response being triggered is dependent on the rate individuals seek testing. In our baseline model, we assume 50% of infected individuals will seek testing, following the incubation period, at a rate such that 99% of these individuals will do so within 8 days. An increase in either the probability of an infected person seeking a test, or the test seeking rate, leads to earlier detection of infection enabling earlier implementation of control strategies (Figure A-2). An increase of 25% in the probability of an infected person seeking a test approximately reduces the number of infections already in the population by the time the first case is identified by half.

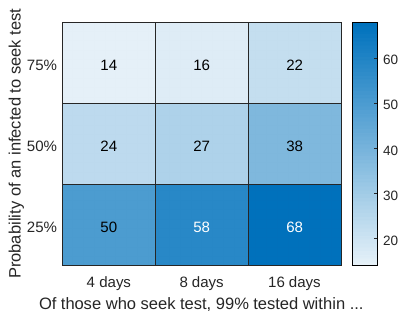

Figure A‑2:The cumulative number of COVID-19 infections in a population of size 1000 by the time the first positive case is identified, as a function of the testing rate and the probability an infected individual will seek a test. Median values from 100 simulations are shown.

##### Proportion of infected individuals that seek testing

In our baseline model we assume that 50% of individuals with SARS-CoV-2 infection will seek testing. This is a broad assumption as at the time of writing there is considerable uncertainty regarding the proportion of SARS-CoV-2 infections that become symptomatic, whether transmissibility varies depending on symptoms, and the likelihood that an infected person will seek testing. To explore the impact of this assumption on outcomes, we ran additional simulations for a representative scenario (isolation + quarantine and testing of all household contacts + clearance test) where either 25% or 75% of infected individuals seek testing.

Outbreaks are initially more difficult to control if the proportion that seek testing is low (Figure A‑3). However, outcomes are similar beyond the initial wave, partly because the majority of subsequent controls, such as contact tracing/testing and lockdown, are applied routinely and thus not affected by the proportion that seeks testing (Figure A‑3).

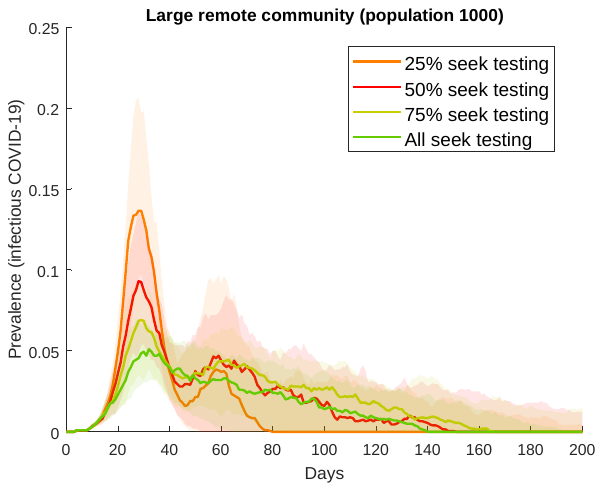

Figure A‑3: Epidemic curves for a population of 1000 in which case is isolated, immediate and extended household members of the case are quarantined and tested, and all individual in isolation/quarantine are subject of clearance testing, with various proportion of infected who seek testing.

##### Extending contact tracing beyond 2 days

In the main text we showed that, in the absence of lockdown, strategies that include the quarantine and testing of a case’s extended household are better able to suppress outbreaks compared to strategies involving the tracing, quarantine and testing of a case’s contacts from the previous two days. We conducted additional simulations to explore the impact of history-based contact tracing over longer periods.

If contacts can be traced beyond 2 days, conventional history-based contact tracing performs better than household-based contact tracing (Figure A‑4 and Table A‑5). There is also a notable reduction in the number of quarantine person days and tests required, since only a subset of an individual’s extended household is included in this strategy.

We note that advice received from relevant health authorities indicates that contact tracing beyond 2 days is not feasible for most remote Aboriginal communities.

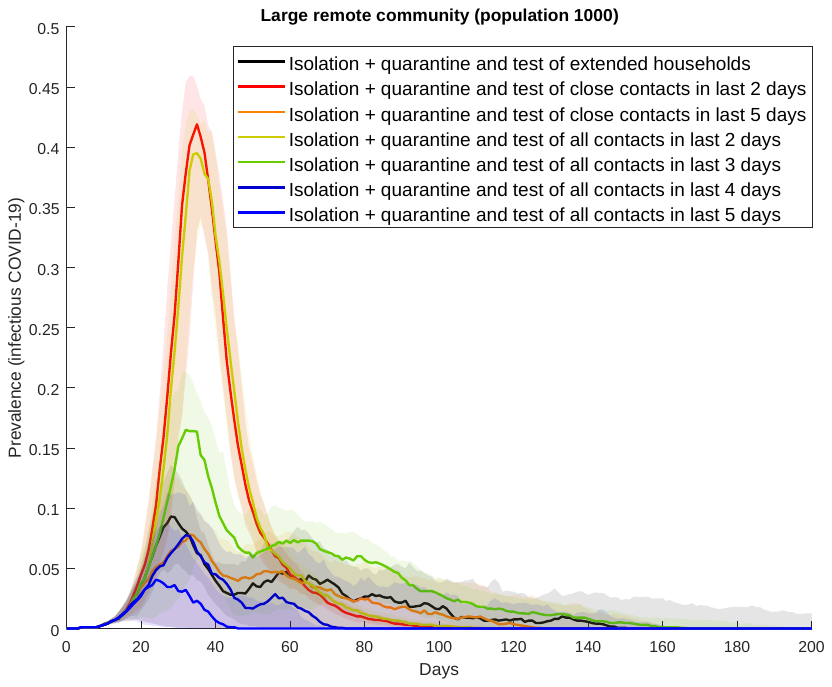

Figure A‑4: Epidemic curves for a response comprising isolation of the case and the quarantine and testing of the case’s household (black), in comparison with isolation of the case and quarantine and testing of the case’s contacts over various periods. Clearance testing is conducted for all those in isolation and quarantine.

##### Supplementary results for selected strategies

Table A‑5: Epidemic characteristics of COVID-19 outbreak in a population of 1000 individuals under selected strategies. The entries are the median outcome from 100 simulations with interquartile range shown in parenthesis.

| **#** | **Strategy** | **Outbreak duration (days)** | **Peak preva-lence (%)** | **Final size of outbreak** | **Number of tests within first year of outbreak** | **Max. number of tests in a single day** | **Number in isolation within first year of outbreak (person-days)** | **Max. number of isolations in a day** | **Number in quaran-tine within first year of outbreak (person-days)** | **Max. number of quaran-tines in a day** |
| --- | --- | --- | --- | --- | --- | --- | --- | --- | --- | --- |
| **0** | No response | 56.0 (54.0, 58.0) | 69.1 (68.0, 70.2) | 999.0 (999.0, 999.0) | 447.0 (435.5, 458.0) | 42.0 (39.5, 45.0) | N/A | N/A | N/A | N/A |
| **1** | Isolation of case | 57.5 (55.5, 60.0) | 67.5 (66.3, 68.4) | 999.0 (999.0, 999.0) | 1221.0 (1188.5, 1250.0) | 53.0 (51.0, 56.0) | 7730.0 (7515.0, 7925.0) | 380.0 (366.0, 388.0) | N/A | N/A |
| **Quarantine** | | | | | | | | | | |
| **2** | #1 + quarantine of immediate households | 69.0 (65.0, 72.0) | 58.3 (56.9, 60.1) | 989.0 (986.0, 991.5) | 9396.0 (9174 - 9572) | 482.5 (463 - 501) | 11850.0 (11633.5 - 12006.5) | 11850.0 (11633.5 - 12006.5) | 2388.5 (2319.0 - 2457.0) | 181.5 (164.0 - 199.0) |
| **3** | #2 + quarantine of extended households | 87.5 (79.0, 99.0) | 45.2 (42.1, 47.6) | 944.0 (934.0, 953.5) | 12012.0 (11713 - 12264) | 521.5 (490 - 549) | 12703.0 (12549.0 - 12879.5) | 554.5 (522.0 - 587.5) | 4577.5 (4327.0 - 4844.5) | 264.5 (236.5 - 285.0) |
| **4** | #1 + quarantine of close contacts from last 2 days | 74.0 (70.0, 80.0) | 52.3 (50.3, 54.3) | 975.0 (967.5, 979.5) | 8827.0 (8665 - 9025) | 409.0 (389 - 433) | 13337.5 (13092.5 - 13516.5) | 627.5 (601.5 - 655.0) | 2430.0 (2359.5 - 2559.0) | 169.0 (152.0 - 187.5) |
| **5** | #4 + quarantine of all contacts from last 2 days | 77.0 (71.0, 82.0) | 51.5 (48.9, 53.2) | 970.0 (964.0, 975.0) | 8937.0 (8786 - 9169) | 417.0 (392 - 439) | 13485.5 (13289.5 - 13612.5) | 630.5 (607.5 - 649.0) | 2635.5 (2463.0 - 2734.5) | 181.5 (156.0 - 199.0) |
| **Entry test on quarantine** | | | | | | | | | | |
| **6** | #2 with test upon entry to quarantine  (Test & quarantine of immediate households) | 141.0 (126.0, 155.0) | 37.6 (31.9, 40.4) | 922.0 (907.5, 936.5) | 1957.5 (1867 - 2027) | 121.0 (102 - 142) | 94.0 (83 - 108) | 116.0 (109.5 - 120.0) | 29595.5 (28101.5 - 31175.0) | 797.5 (762.5 - 825.0) |
| **7** | #3 with test upon entry to quarantine  (Test & quarantine of extended households) | 242.0 (155.0, 292.0) | 13.7 (10.7, 16.5) | 831.5 (751.0, 871.0) | 4042.5 (3463 - 4305) | 206.5 (159 - 257) | 3027.5 (2332.0 - 3306.5) | 31.5 (29.0 - 33.5) | 86825.0 (70334.5 - 97662.5) | 912.5 (867.0 - 936.0) |
| **8** | #4 with test upon entry to quarantine  (Test & quarantine of close contacts) | 102.5 (92.5, 111.5) | 45.7 (43.1, 48.1) | 937.0 (929.0, 945.0) | 1530.5 (1441 - 1586) | 92.5 (83 - 105) | 10981.5 (10805.5 - 11118.5) | 522.0 (489.5 - 555.5) | 10776.5 (9551.5 - 11564.5) | 303.0 (289.0 - 316.5) |
| **9** | #5 with test upon entry to quarantine  (Test & quarantine of all contacts) | 109.0 (95.0, 120.5) | 43.8 (40.9, 46.4) | 930.0 (917.0, 941.0) | 1614.5 (1550 - 1667) | 94.0 (83 - 108) | 11025.5 (10837.0 - 11238.0) | 510.5 (469.0 - 544.0) | 11887.0 (11180.0 - 12831.5) | 322.0 (306.0 - 334.5) |
| **Entry and clearance test on quarantine** | | | | | | | | | | |
| **10** | #6 with clearance test upon exit from quarantine  (Test & quarantine of immediate households + clearance test) | 136.5 (126.5, 154.5) | 36.5 (31.8, 40.7) | 922.5 (905.0, 933.0) | 7526.0 (7336 - 7743) | 319.0 (268 - 371) | 8689.5 (8392.5 - 8942.5) | 294.0 (260.0 - 317.5) | 22500.5 (21469.0 - 23306.0) | 743.0 (683.0 - 771.5) |
| **11** | #7 with clearance test upon exit from quarantine  (Test & quarantine of extended households + clearance test) | 115.5 (49.0, 238.0) | 13.2 (10.6, 15.3) | 655.0 (267.5, 821.0) | 13551.5 (4929.5, 16729.5) | 474.0 (395.5, 581.5) | 6218.5 (2277.5, 7576.5) | 110.5 (89.0, 126.5) | 50958.0 (13511.5, 67786.0) | 841.5 (752.0, 908.5) |
| **12** | #8 with clearance test upon exit from quarantine  (Test & quarantine of close contacts + clearance test) | 102.0 (93.5, 115.0) | 46.0 (42.8, 48.1) | 930.5 (917.0, 939.5) | 4673.5 (4549.5, 4780.5) | 178.5 (168.5, 191.0) | 14265.5 (14040.0, 14658.5) | 570.0 (529.0, 597.0) | 9445.5 (8541.5, 10191.5) | 300.0 (282.0, 317.0) |
| **13** | #9 with clearance test upon exit from quarantine  (Test & quarantine of all contacts + clearance test) | 107.5 (92.5, 123.0) | 43.3 (40.2, 46.1) | 919.0 (904.5, 931.5) | 4842.5 (4741.0, 4957.0) | 181.0 (165.0, 199.0) | 14373.5 (14080.0, 14587.5) | 551.0 (518.5, 579.0) | 10662.0 (9718.0, 11768.5) | 317.5 (303.0, 339.0) |
| **Lockdown** | | | | | | | | | | |
| **14** | #7 + lockdown  (Test & quarantine of extended households + lockdown) | 238.0 (38.5, 278.3) | 12.6 (9.7, 15.6) | 829.0 (712.0, 866.5) | 3927.5 (3434.5, 4156.0) | 190.5 (139.0, 250.0) | 2909.0 (2297.0, 3189.5) | 31.0 (28.0, 33.0) | 85283.0 (69397.0, 92022.5) | 894.0 (818.0 - 929.5) |
| **15** | #11 + lockdown  (Test & quarantine of extended households + clearance test for quarantine + lockdown) | 41.0 (34.3, 72.5) | 6.5 (1.6, 12.4) | 88.5 (20.0, 432.5) | 1402.0 (344.5, 7564.0) | 198.0 (67.0, 432.0) | 752.0 (197.0, 3703.0) | 45.5 (11.5, 103.5) | 5253.5 (1660.5, 24531.0) | 384.0 (118.0, 776.0) |
| **16** | #11 + lockdown + clearance test from lockdown  (Test & quarantine of extended households + lockdown + clearance test for quarantine & lockdown) | 35.5 (29.5, 39.0) | 3.1 (0.9, 5.2) | 35.0 (9.0, 62.5) | 2498.0 (2169.5, 2823.5) | 1000.0 (1000.0, 1000.0) | 420.5 (128.0, 735.5) | 29.5 (8.0, 52.5) | 3469.0 (1431.5, 5602.5) | 286.0 (112.0, 452.5) |
| **Delays between identification of cases and intervention** | | | | | | | | | | |
|  | #11  (Test & quarantine of extended households + clearance test, with delay of 2 days) | 115.5 (49.0, 238.0) | 13.2 (10.6, 15.3) | 655.0 (267.5, 821.0) | 13551.5 (4929.5, 16729.5) | 474.0 (395.5, 581.5) | 6218.5 (2277.5, 7576.5) | 110.5 (89.0, 126.5) | 50958.0 (13511.5, 67786.0) | 841.5 (752.0, 908.5) |
| **17** | #11 with delay of 4 days  (Test & quarantine of extended households + clearance test) | 54.0 (50.5, 89.0) | 27.4 (23.2, 33.5) | 486.0 (403.0, 612.5) | 7547.0 (6997 - 10853) | 7547.0 (6997 - 10853) | 3902.0 (3354.5 - 5345.0) | 244.0 (203.5 - 300.0) | 14078.5 (13633.0 - 27743.5) | 953.0 (931.0 - 964.0) |
| **18** | #11 with delay of 6 days  (Test & quarantine of extended households + clearance test) | 56.0 (54.0, 95.0) | 46.3 (40.7, 52.2) | 752.5 (678.5, 839.0) | 11239.0 (10290 - 13318) | 898.5 (866 - 927) | 5899.5 (5499.5 - 6826.5) | 5899.5 (5499.5 - 6826.5) | 953.0 (931.0 - 964.0) | 963.0 (953.0 - 968.0) |
|  | #16 (Test & quarantine of extended households + lockdown + clearance test for quarantine & lockdown, with delay of 2 days) | 35.5 (29.5, 39.0) | 3.1 (0.9, 5.2) | 35.0 (9.0, 62.5) | 2498.0 (2169.5, 2823.5) | 1000.0 (1000.0, 1000.0) | 420.5 (128.0, 735.5) | 29.5 (8.0, 52.5) | 3469.0 (1431.5, 5602.5) | 286.0 (112.0, 452.5) |
| **19** | #16 with delay of 4 days  (Test & quarantine of extended households + clearance test) | 41.0 (36.5, 44.0) | 7.7 (3.0, 12.9) | 92.0 (34.5, 164.0) | 3059.0 (2404 - 3659) | 1000.0 (1000 - 1000) | 1236.0 (494.0 - 2088.5) | 85.0 (32.5 - 148.5) | 4694.0 (2121.0 - 6797.5) | 452.0 (201.5 - 653.0) |
| **20** | #16 with delay of 6 days  (Test & quarantine of extended households + clearance test) | 46.0 (41.0, 50.0) | 15.4 (7.4, 29.4) | 204.5 (96.5, 397.0) | 3658.5 (2889 - 4693) | 1000.0 (1000 - 1000) | 3138.5 (1512.0 - 5630.5) | 199.0 (94.5 - 379.0) | 5689.0 (3309.5 - 7515.5) | 660.0 (379.0 - 877.0) |
| **Compliance with lockdown** | | | | | | | | | | |
| **21** | #16 with 50% compliance to lockdown  (Test & quarantine of extended households + lockdown + clearance test for quarantine & lockdown) | 76.5 (33.0, 245.0) | 11.3 (1.3, 14.3) | 415.5 (18.0, 823.5) | 9490.5 (1463 - 17529) | 518.0 (503 - 554) | 4047.5 (255.0 - 7661.5) | 98.0 (14.0 - 119.0) | 28117.5 (2535.5 - 67538.5) | 765.0 (198.0 - 856.0) |
| **22** | #16 with 60% compliance to lockdown  (Test & quarantine of extended households + lockdown + clearance test for quarantine & lockdown) | 63.0 (30.0, 160.8) | 11.4 (1.3, 14.5) | 280.5 (15.0, 689.5) | 6705.5 (1592 - 14587) | 609.5 (601 - 623) | 2563.0 (213.0 - 6264.5) | 102.0 (12.0 - 118.5) | 17748.0 (2086.0 - 46952.0) | 792.0 (164.0 - 865.5) |
| **23** | #16 with 70% compliance to lockdown  (Test & quarantine of extended households + lockdown + clearance test for quarantine & lockdown) | 64.5 (29.0, 211.0) | 10.0 (1.1, 14.5) | 331.5 (11.0, 794.0) | 7174.5 (1678 - 16866) | 706.0 (701 - 714) | 3112.5 (155.5 - 7495.5) | 89.0 (10.0 - 115.5) | 16898.0 (1702.0 - 62568.0) | 721.0 (135.5 - 848.0) |
| **24** | #16 with 80% compliance to lockdown  (Test & quarantine of extended households + lockdown + clearance test for quarantine & lockdown) | 40.0 (30.0, 189.5) | 4.3 (1.0, 12.4) | 57.0 (11.5, 769.5) | 2542.0 (1840 - 16947) | 804.0 (801 - 809) | 660.0 (140.0 - 7311.0) | 45.0 (9.5 - 106.0) | 4991.0 (1635.5 - 60108.0) | 400.5 (128.5 - 814.0) |
| **25** | #16 with 90% compliance to lockdown  (Test & quarantine of extended households + lockdown + clearance test for quarantine & lockdown) | 37.0 (28.0, 44.5) | 3.2 (1.2, 10.4) | 38.0 (14.0, 191.0) | 2409.0 (2083 - 4025) | 902.5 (900 - 905) | 454.5 (182.5 - 1874.5) | 31.0 (11.5 - 89.5) | 3718.5 (1581.5 - 10021.5) | 310.0 (129.0 - 728.0) |
| **Proportion of infected individuals that seek testing** | | | | | | | | | | |
| **26** | #11 with 25% infection seek testing  (Test & quarantine of extended households + clearance test) | 74.5 (49.0, 211.5) | 19.4 (15.6, 24.0) | 508.0 (310.5, 877.5) | 8829.5 (5129 - 15640) | 689.0 (585 - 771) | 4176.0 (2520.0 - 7285.0) | 153.0 (125.0 - 192.5) | 25679.5 (13325.5 - 60818.5) | 936.5 (906.5 - 957.0) |
|  | #11  (Test & quarantine of extended households + clearance test, 50% infection seek testing) | 115.5 (49.0, 238.0) | 13.2 (10.6, 15.3) | 655.0 (267.5, 821.0) | 13551.5 (4929.5, 16729.5) | 474.0 (395.5, 581.5) | 6218.5 (2277.5, 7576.5) | 110.5 (89.0, 126.5) | 50958.0 (13511.5, 67786.0) | 841.5 (752.0, 908.5) |
| **27** | #11 with 75% infection seek testing  (Test & quarantine of extended households + clearance test) | 151.5 (57.0, 234.0) | 10.3 (8.3, 12.4) | 586.5 (296.5, 742.5) | 13130.5 (6862 - 16945) | 364.0 (301 - 437) | 5843.5 (2799.0 - 7406.5) | 90.0 (77.0 - 108.0) | 49583.0 (24754.0 - 69079.0) | 767.0 (705.5 - 846.0) |
| **28** | #11 with all infection seek testing  (Test & quarantine of extended households + clearance test) | 138.5 (54.0, 188.0) | 8.0 (6.5, 9.4) | 449.0 (160.0, 594.0) | 10842.5 (4187 - 13909) | 299.5 (246 - 367) | 4794.5 (1662.0 - 6199.5) | 74.0 (62.0 - 84.0) | 41500.0 (15293.0 - 56783.5) | 686.0 (600.0 - 745.0) |
| **Extended contact history** | | | | | | | | | | |
| **29** | #1 + quarantine and test of all case’s contacts from last 3 days + clearance test | 164.5 (39.0, 195.0) | 20.1 (14.3, 23.5) | 845.5 (749.0, 886.0) | 7858.0 (6882.5, 8420.5) | 190.0 (145.5, 217.0) | 11736.0 (10773.0, 12191.0) | 232.5 (183.5, 259.0) | 33046.5 (25833.0, 36931.0) | 582.5 (448.0, 640.5) |
| **30** | #1 + quarantine and test of all case’s contacts from last 4 days + clearance test | 71.5 (29.0, 163.5) | 11.5 (1.0, 14.9) | 374.5 (10.0, 671.0) | 4873.5 (204.0, 9001.5) | 205.0 (54.5, 242.5) | 5031.5 (171.0, 8731.5) | 132.0 (9.0, 151.0) | 19383.5 (1021.0, 41243.5) | 619.0 (77.0, 703.0) |
| **31** | #1 + quarantine and test of all case’s contacts from last 5 days + clearance test | 45.5 (28.0, 68.0) | 6.8 (0.8, 10.8) | 133.0 (8.0, 262.0) | 2329.0 (210.5, 4453.5) | 194.5 (53.5, 274.0) | 1625.0 (140.5, 3162.5) | 71.5 (7.0, 106.5) | 10132.0 (1004.5, 18404.5) | 542.5 (75.0, 710.5) |
| **32** | #1 + quarantine and test of case’s close contacts from last 5 days + clearance test | 115.5 (32.0, 164.5) | 11.8 (1.0, 14.5) | 544.0 (11.0, 731.5) | 7509.0 (276.5, 9850.0) | 223.0 (69.5, 267.0) | 6538.0 (162.0, 8628.0) | 117.5 (9.0, 143.0) | 32250.5 (1407.5, 43749.0) | 642.5 (106.0, 717.5) |

13.	Australian Bureau of Statistics: **General Social Survey, Summary Results, Australia, 2014**. In*.* Edited by Australian Bureau of Statistics; 2015.
